# VEGF subtype A and B Gene Expression, Clues to a Temporal Signature in Kawasaki Disease, Implications for Coronary Pathogenesis through a Secondary analysis of Clinical Datasets

**DOI:** 10.1101/2022.08.08.22278559

**Authors:** Asrar Rashid, M. Toufiq, Praveen Khilnani, Zainab A. Malik, Javed Sharief, Raziya Kadwa, Berit S. Brusletto, Amrita Sarpal, Damien Chaussabel, Rayaz Malik, Nasir Quraishi, Govind Benakatti, Syed A. Zaki, Pauline Ogrodzki, Walid Zaher, Mouhamad Al Zouhbi, Husam Saleh, Ahmad Al Khayer, M. Rashid Nadeem, Guftar Shaikh, Shazia Hussain, Mishal Tariq, Ahmed Al-Dubai, Amir Hussain

## Abstract

**Background and Aim:** An essential issue for Kawasaki Disease (KD) is the development of coronary artery disease. We decided to investigate (VEGF) subtype gene expression in KD due to the proangiogenic nature of Vasoactive Endothelial Growth Factor A (VEGF). VEGF-A is a known angiogenic molecule with pro-inflammatory effects, whereas the role of VEGFB has been less defined.

**Method:** KD Microarray and RNA-seq datasets were selected using a comprehensive search strategy of the NCBI GEO Dataset, which resulted in eight studies from whole blood. This included three extensive studies in KD (KD1-KD3). Further, one study dataset from coronary artery tissue, the Coronary Artery Dataset (CAD), was also included to appreciate end-stage KD.

**Results:** In CAD, cases of KD versus controls, VEGF-B was up-regulated (p = 4.932e-02). KD1, KD Acute versus convalescent samples, VEGF-A is up-regulated (p=1.258e-07) and VEGF-B was down-regulated (p=1.42e-28). Similar up regulation of VEGF-A and down regulation of VEGF-B was seen in KD2 (p=1.140e-04; p=1.746e-02) and KD3 (p=1.140e-04; p=1.746e-02), both are KD Versus Controls. VEGF-A up-regulated (p=1.140e-04), VEGF-B down-regulated (p=1.746e-02).KD3, KD versus Control; VEGF-A up-regulated (p=1.140e-04), VEGF-B down regulated (p=1.746e-02).

**Conclusions:** In acute KD VEGF-A up-regulation, with VEGFB being down-regulated, the reverse being true of the convalescent situation. This suggests a temporal inverse relationship between VEGF-A and VEGF-B may have biomarker implications. Moreover, a dual therapeutic strategy, enhancing VEGFB while minimizing VEGFA effects, could be a possible advancement over current KD related-therapies. Further work defining the relationship of VEGF-A to VEGF-B in KD-related angiogenesis is suggested.

## Introduction

Kawasaki Disease (KD) remains a leading cause of acquired childhood heart disease in developing nations, impacting medium-sized vessels. Further, a KD sub-phenotype of Multi-Inflammatory Syndrome in children (KD MIS-C) associated with the SARS-CoV-2 pandemic has been described. In both KD and KD MIS-C, the worrisome complication relates to coronary artery disease, thus the need to focus on the question of coronary artery pathogenesis in KD. Accordingly, a potent angiogenic factor, Vasoactive Endothelial Growth Factor (VEGF), has been investigated in the context of KD. VEGF levels are increased in the acute stage in children without coronary pathology and in the sub-acute phase in children with coronary pathology {Maeno, 1998 #2795}. Moreover, Maeno et al. showed a temporal difference in VEGF expression in neutrophils versus peripheral blood mononuclear cells (PBMCs). Thereby suggesting that VEGF-A could be part of an early response leading to coronary artery abnormalities. This is consistent with the finding of Maeno et al. {Maeno, 1998 #2374} who found elevated VEGF-A in the acute phase of KD, with yet a further elevation of VEGF-A in the subacute stage. One possible explanation as to why coronary artery abnormalities occur less often in adults could be due to lowering endothelial responsiveness to VEGF-associated angiogenesis, noted with advancing age {Ungvari, 2018 #2390}.

The parent molecule, VEGF, is an angiogenic factor consisting of different mammalian homologs, including VEGF-A, B, C, D, and Placenta Growth Factor (PGF). VEGF-A has physiological and pathological regulations affecting angiogenesis and vasculogenesis {Eguchi, 2022 #2921}. Further, Breunis et al. showed that VEGF-A gene polymorphisms may play a role in KD pathogenesis {Breunis, 2006 #2396}. Hamachi et al. showed that KD patients developed coronary artery changes associated with increased serum VEGF-A throughout the 2 to 4 weeks after the onset of KD illness {Hamamichi, 2001 #2924}. Showing VEGF-A to be driven from Peripheral Blood Mononuclear Cells (PBMC) through to the subacute stage of the KD illness. Thus providing further evidence for the importance of VEGF-A in KD pathogenesis.

Also VEGF-A is known to be pro-inflammatory, but its role in KD coronary artery disease and its relationship to the KD remains elusive. Huang et al. found increased VEGF-A as well as VEGF receptor type 2 (VEGFR2) associated with increased Endothelial Cell (EC) permeability {Huang, 2021 #2920}. VEGF molecules bind to VEGFR receptors (VEGFR), including subtypes 1 to 3 and neurolipins (subtypes 1 and 2). VEGF-A biding to VEGFR2 promotes the strongest impulse to angiogenesis compared to other VEGF-VEGFR or -neuropilin pairings{Ferrara, 2004 #2922}. Unlike VEGF-A, VEGF-B only binds to VEGFR-1{Nash, 2006 #2924}. VEGF-C and D are non-angiogenic, influencing lymphangiogenesis instead. Further, it is known that VEGF B alters gene expression of proteins involved in cardiac metabolism and promotes cell survival {Lal, 2018 #2787}. The fact that VEGF-B, unlike VEGF-A, does not induce neovascularisation under most circumstances is perplexing {Arjunan, 2018 #3149}. However, VEGF-B is found to be critical for vascular survival, also seeming dispensable for blood vessel growth {Zhang, 2009 #2923}. However, Tomek et al. demonstrated dose-dependent VEGF-B inhibition resulting in the limitation on coronary tube formation{Tomanek, 2002 #3151}. Moreover, literature regarding the role of VEGF-B in KD disease pathogenesis is sparse, unlike VEGF-A, which has been extensively studied. Therefore, the pathophysiological role of both VEGF-A & VEGF-B (VEGF-A/B) and pathophysiological consequences in KD require further attention.

There is a desperate clinical need for a biomarker to improve the precision of KD treatment, well before echocardiographic findings become suggestive of coronary artery changes. Ideally, such a KD biomarker should improve predictability and allow immunological signposting, i.e., to demarcate different phases of the illness. We postulate that gene differential expression patterns of VEGF-A/B subtypes in KD could provide invaluable insights in terms of biomarker opportunities. The fact that whole blood gene expression KD studies may not indicate changes at the coronary artery level is challenging, given the accessibility of whole blood. To understand this predicament, a coronary artery study by Rowley et al. is included as a proxy for end-stage KD {Rowley, 2015 #2394}.

## Material and Methods

### Systematic Dataset Search Strategy and Data Pre-Processing

In the present study, datasets from the NCBI GEO datasets (http://www.ncbi.nlm.nih.gov/geo) were searched using the keywords ‘Kawasaki Disease’ [MeSH Terms] [All Fields] AND ‘Homo sapiens’ [porgn] AND ‘gse’ [Filter]. Only data from RNA-Seq or Microarray studies were included. Further non-random selection was carried out based on; 1) the comparison of genomic and transcriptome profiles between KD patients, patients with bacterial or viral infection and healthy controls, 2) KD patients and control with coronary arteritis, and 3) KD treatment studies. A total of ten datasets were identified using a systematic search strategy (Figure 1). Datasets included GSE63881, GSE73461, GSE68004, GSE64486, GSE16797, GSE15297, GSE109351, GSE48498 and GSE18606 were included in the final analysis (Table 1). However, GSE73577 was excluded due to the inability to classify the dataset by virtue of inadequate labeling of samples. The datasets were first inspected for data processing methods as indicated by the original study author(s). The appropriate normalization and log2 transformation were applied as needed using R-script for further downstream analysis. The KD datasets then underwent a comprehensive program of gene and pathway enrichment. The aim was to specifically focus on VEGF-A and VEGF-B gene expression in order to understand their role in KD.

**Figure 1.**
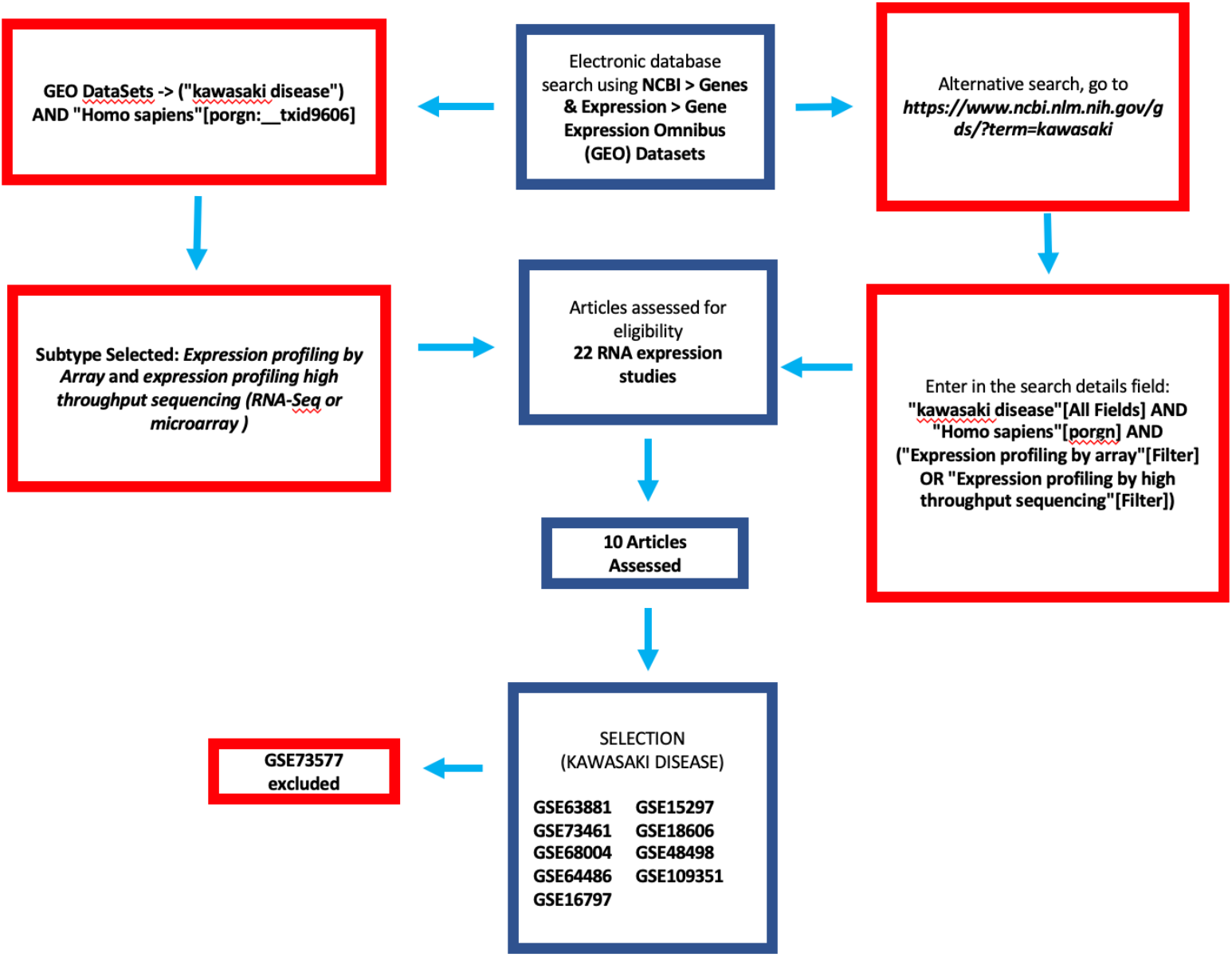
Systematic Search for KD datasets from DNA Microarray and RNA-Seq datasets. Discriminate Molecular Mechanisms between KD positive and the Controls. In the present study, datasets from the GEO database (http://www.ncbi.nlm.nih.gov/geo) were searched using the key words ‘Kawasaki Disease’ [MeSH Terms] [All Fields] AND ‘Homo sapiens’ [porgn] AND ‘gse’ [Filter]. Only data from RNA-Seq or microarray studies were included. Further non-random selection was carried out based on; 1) the comparison of genomic and transcriptome profiles between KD patients, patients with bacterial or viral infection and healthy controls, 2) KD patients and control with coronary arteritis, and 3) KD treatment and controls. A total of ten datasets, GSE63881, GSE73461, GSE68004, GSE64486, GSE16797, GSE15297, GSE109351, GSE48498 and GSE18606 were included in the final analysis (Table 1). However, GSE73577 was excluded due to insufficient labelling of the dataset. All microarray datasets were pre-processed by log2 quantile normalisation using R script.

**TABLE 1:**
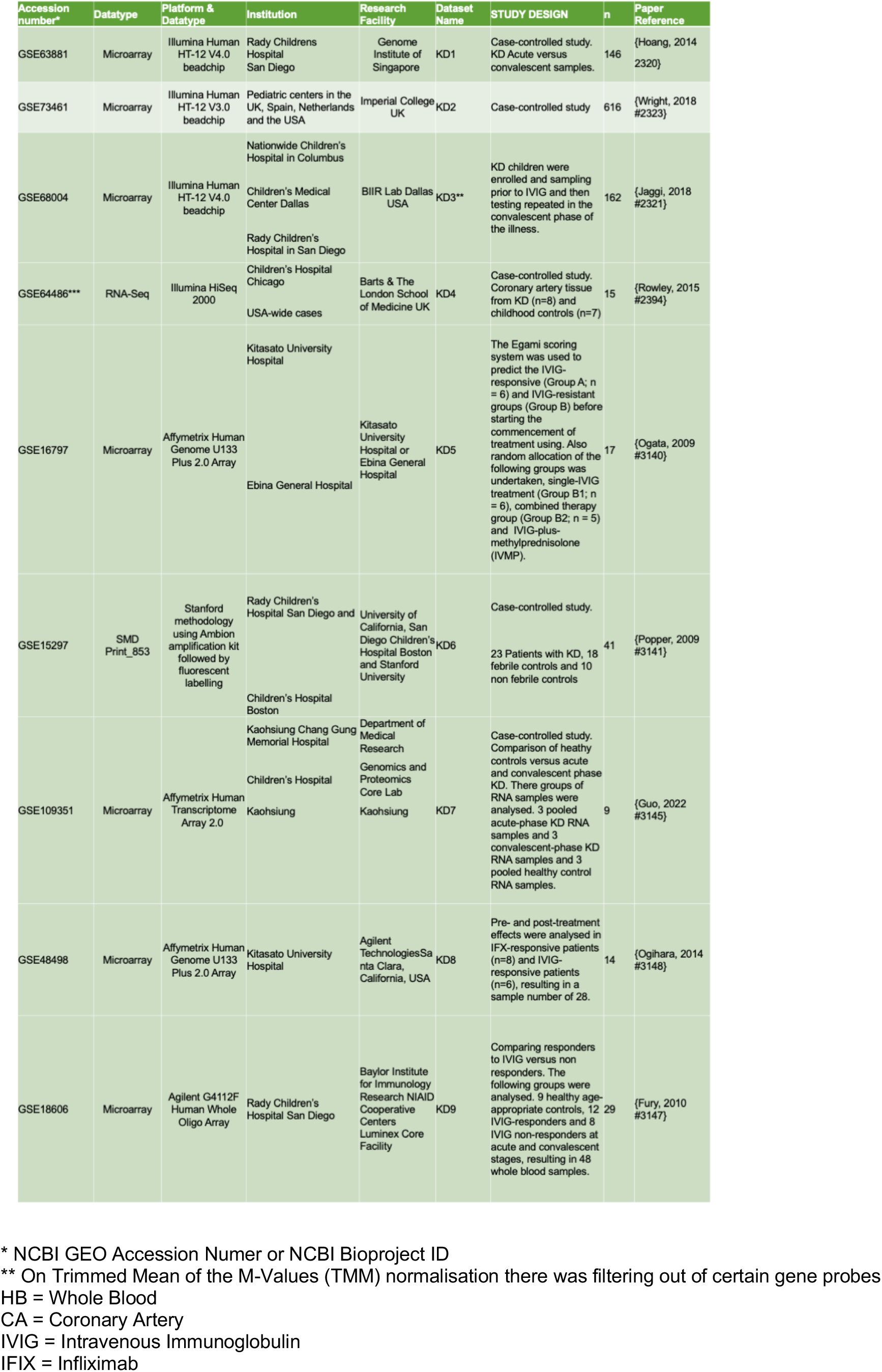
Summary of KD Clinical Studies Analysed.

### Statistical and Gene Ontology analyses

The aim was to analyze KD clinical studies using the Differential gene expression (DGE) on the Qlucore Omics Explorer (QOE) version 3.1 software platform (Qlucore AB, Lund, Sweden). Principal Component Analysis (PCA) plots were generated using QOE. Thus allowing two group and multi-group comparison as well as unsupervised hierarchical clustering. mRNA array data was analyzed with Qlucore Omics Explorer version 3.7 for DGE. Every probe in the expression file was allocated to its respective HGNC (HUGO Gene Nomenclature Committee) [26842383] Gene Symbol(s). Duplicate “Gene Symbol(s)” mapping to multiple probe IDs was collapsed by averaging. In order to understand the genes of interest, the QOE Gene Symbols setting was used followed by probe averaging. Hierarchical clustering was based on Euclidean distance and average linkage clustering. Further, all genes were centered to mean equal to zero and scaled to variance equal to one. Value q, also known as the False discovery rate (FDR) was deemed statistically significant if its value was below 0.25. QOE was also used to generate a PCA heat plot of genes of interest. Various gene enrichment techniques were used through software solutions ShinyGO version 0.76 {Ge, 2020 #3213}, g-profiler version 2020 {Reimand, 2016 #2100} and GSEA provided through QOE 3.1 {Subramanian, 2005 #2600}{Mootha, 2003 #2179}.

### Gene Set Enrichment Analysis

Pathway enrichment analysis was undertaken using Gene Set Enrichment Analysis (GSEA) through the QOE platform. Gene sets were downloaded from the MSigDB database according to terms representing biological molecules. Gene symbol was set at the average setting, prior to importing into the QOE GSEA. For GSEA the set p-value < 0.05 and q value <0.25 was defined as significant. Also, a Net Enrichment Score (NES) was calculated for each gene set. Thereby a highly positive NES implies an upregulated gene set and a high negative NES suggests a downregulated gene set. Permutations are calculated to estimate the p-value and then in order to account for differences in gene sets size and normalization is performed to generate the NES. Query gene sets are used from the collection of gene sets from the Molecular Signature Database (MSigDB, https://gsea-msigdb.org/gsea/msigdb/collections.jsp){Subramanian, 2005 #2178}.

### Enrichment analysis using the ShinyGO platform

The ShinyGo (version 0.76) was also used for gene enrichment analysis. The software resides on the webserver (http://bioinformatics.sdstate.edu/go/) {Ge, 2020 #3213}. Shiny application consists of several R/Bioconductor packages. Default settings are used with a background for all protein folding genes in the genome with an FDR cut-off of 0.05 mapping onto the Gene Ontology (GO) Biological processes database and to the Kyoto Encyclopedia of Genes and Genomes (KEGG) database enabling biological interpretation. Query genes are mapped to a database of gene IDs enabling ID conversion. The features used in this study from the ShinyGO software platform include the ability to visualize enriched genes on pathway diagrams, PPI networks, hierarchical clustering, and interactive networks. Also, ShinyGo has an application program interface (API) with access to KEGG and STRING. This then allows ShinyGO to generate KEGG pathway diagrams.

### Analyzing Angiogenesis from Whole blood and Coronary Artery Tissue

GSE64486 dataset, containing RNA-seq data generated from coronary artery tissue from Kawasaki Disease patients and controls. Herein, 8 KD coronary artery samples were compared to 7 childhood coronary artery controls (non KD). 7 of the 8 KD coronary arteries were post-mortem samples, one being from a child who received a heart transplant. Given the type of samples, the Rowley data-set was assumed to represent end-stage KD. Initially, the paired-end RNA-seq FASTQ files for each sample within the dataset GSE64486 were obtained using the tool parallel-fastq-dump (version 0.67, fastq-dump version 2.8.0). The reads were then mapped to the human reference genome (hg38 obtained from UCSC, URL: https://hgdownload.soe.ucsc.edu/goldenPath/hg38/bigZips/hg38.fa.gz) using the BWA-MEM algorithm (bwa version 0.7.17-r1188). Finally, the resulting SAM files, containing the mapped reads, were sorted by coordinate using Picard - SortSam (version 2.25.5). Thus, the coordinate-sorted mapped BAM files were obtained per sample. The BAM files were then imported into QOE. A GTF (Gene Transfer Format) file is required to understand where genes are located on the reference genome. A Gene Transfer Format (GTF) file was generated from the UCSC.edu table browser using the settings class ‘Mammal’, genome ‘Human’ assembly Dec.2013 (GRCh38/hg38), group ‘Genes and Gene Predictions’, track ’Gencode V36’, table ‘knownGene”. BAM files thus imported into QOE were then normalized in the Trimmed Mean of the M-Values (TMM) normalization prior to further analysis.

## Results

### Coronary Arteritis (KD4: GSE64486)

Analysis of coronary KD4 is shown (Figure 2). Here, VEGF-B gene expression was down-regulated in KD patients versus controls (p = 4.932e-02) with VEGF subtypes (A, C, and D) showing no significant difference, which was also the case for TNF and NFKB1 gene expression. (Figure 2 A-D).

**Figure 2.**
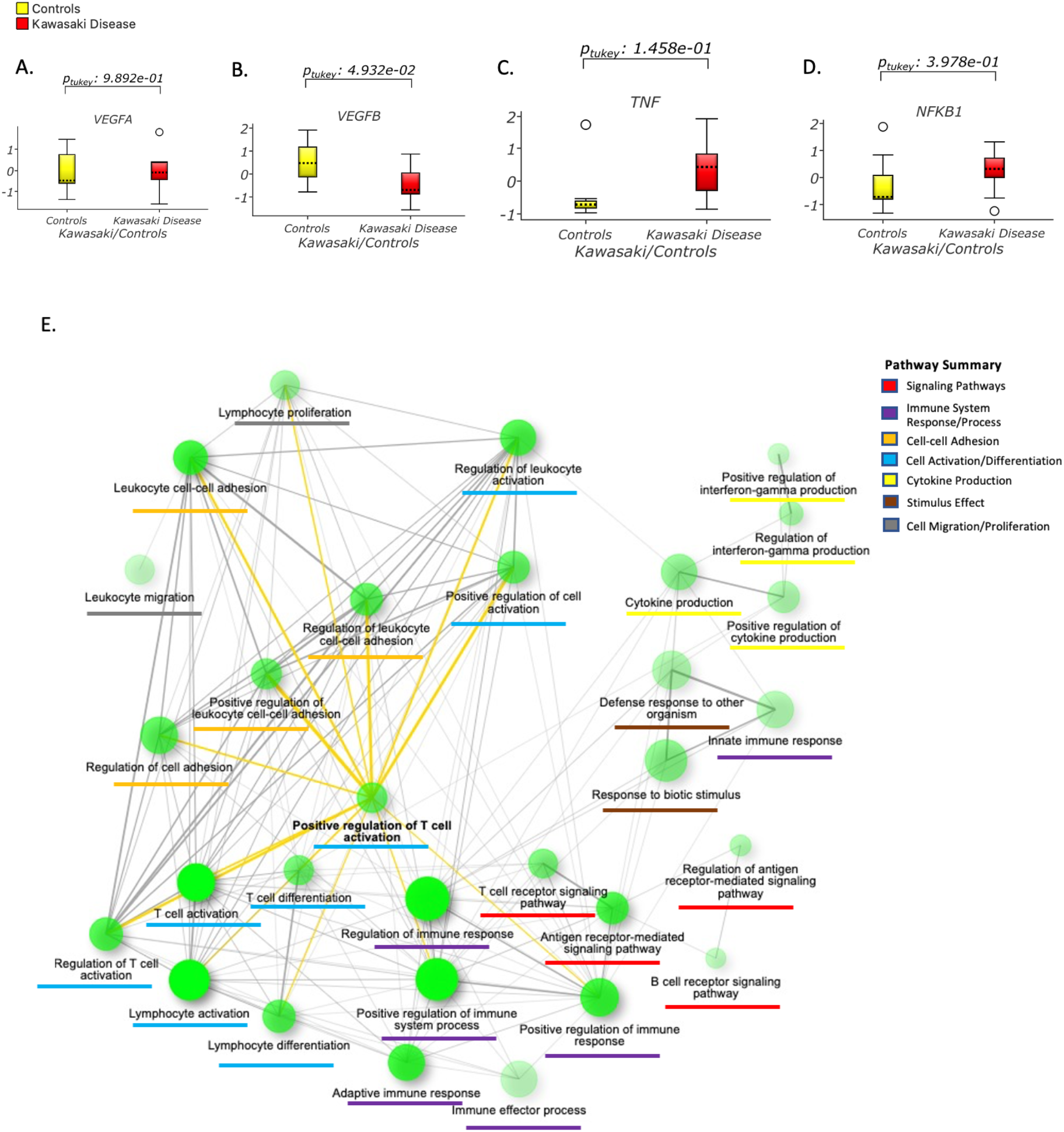
Coronary arteritis in Kawasaki disease compared to controlled patients from deceased samples. Using the childhood coronary artery dataset **KD4** (GSE64486). Gene expression box plots are shown **(A-D)**, only VEGFB significantly up-regulated compared to controls. t-Test Kawasaki Disease versus control (p=0.001, q=0.12) elicited 527 genes which then underwent enrichment and the GO network generated. The SHINYGO network enrichment pattern is shown **(E)** illustrating the GO terms as shown, including antigen receptor-mediated signaling pathway, lymphocyte and leucocyte activation as well as others.

GSEA of the KD4 dataset using t-Test comparison of KD versus controls led genesets of significance as listed (Table 1). GO pathways were elicited including those related to leukocyte adhesion, vascular endothelial cells, and tumor necrosis factor binding. GO positive regulation pathways for interleukin 1, 6, and 8 production, and interferon-gamma production were also noted (Table 2).

**Table 2.**
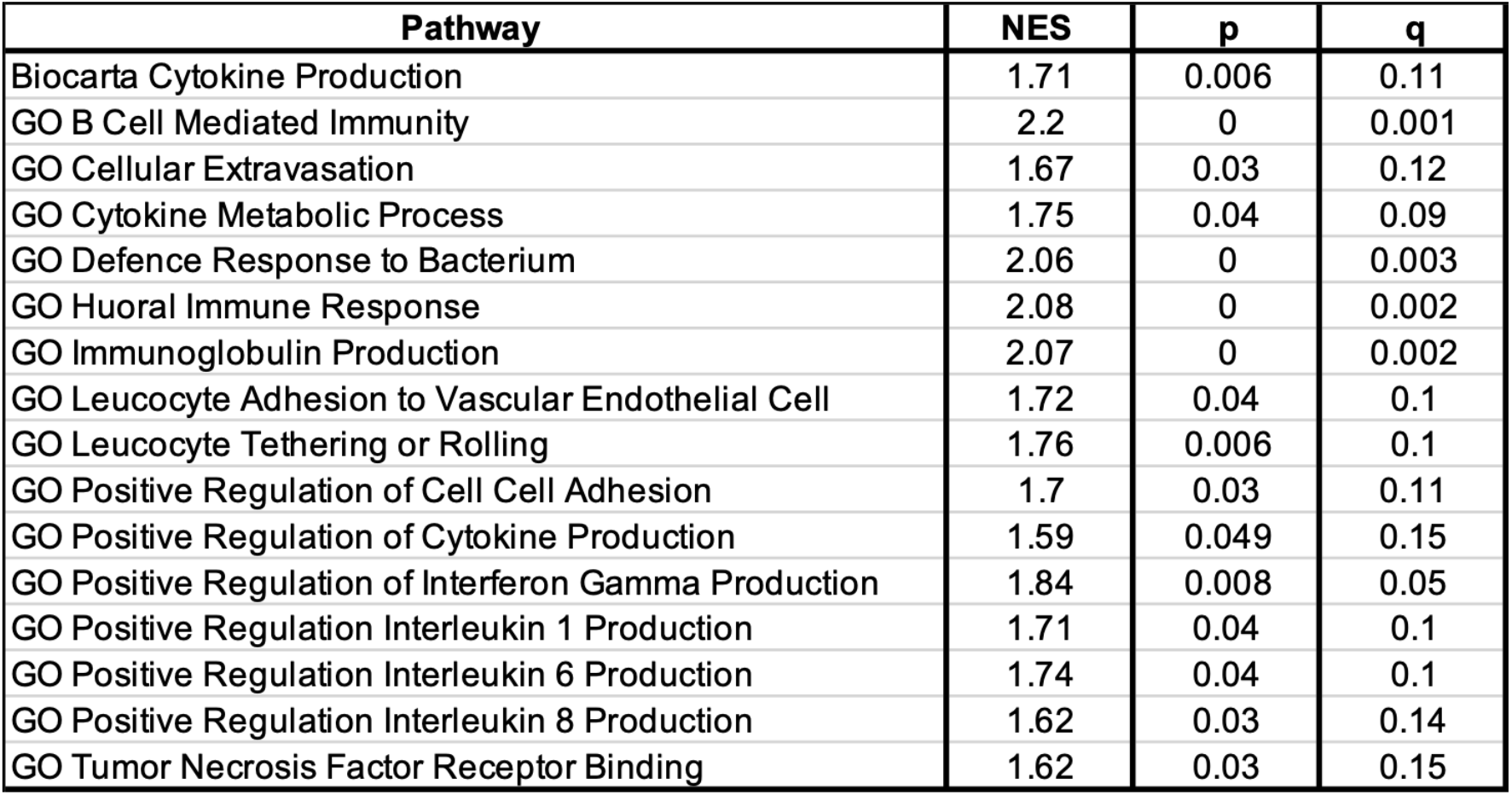
Coronary Artery Gene Set Expression analysis (GSEA). Coronary artery dataset KD4 (GSE 64486) underwent GSEA of Cases versus controls. Variables were identified according to the averaging of their Genes Symbols. A two group comparison was undertaken p<0.05 and q <0.25. Elicited GO pathways are shown. Gene set used was TNF.gmt for GSEA.

Various innate and adaptive immune responses are outlined according to GO pathways (Supplement Table 3A, B, and C).

### PCA Heat Maps Responses

#### Kawasaki Disease - VEGF gene relationships

Two KD datasets showed increased VEGF-A and decreased VEGF-B in acute versus convalescent samples (KD1, Figure 3A & D), and KD versus controls (KD2, Figure 3B & E). In KD3 VEGF-B gene expression was down-regulated for children with KD versus controls, whereas VEGF-A gene expression was unregulated in children with KD versus controls (KD3, Figure 3C & F). Only in KD1 was VEGF-C up-regulated in acute versus convalescent samples. KD5 showed no differences in pre-treatment and post-treatment VEGF-A or VEGF-B, but there was a significant fall in gene expression pre-treatment to post-treatment for NFKB1 (ptukey 6.745e-03) (Figure 5A Supplement). KD6 no difference between febrile patients and those with KD for VEGF-A, VEGF-B, TNF, and NFKB1 (Figure 5B Supplement). KD7 no difference was noted for KD acute phase versus convalescent-phase versus controls for VEGF-A, VEGF-B, TNF, and NFKB1 gene expression (Figure 5C Supplement).

**Figure 3.**
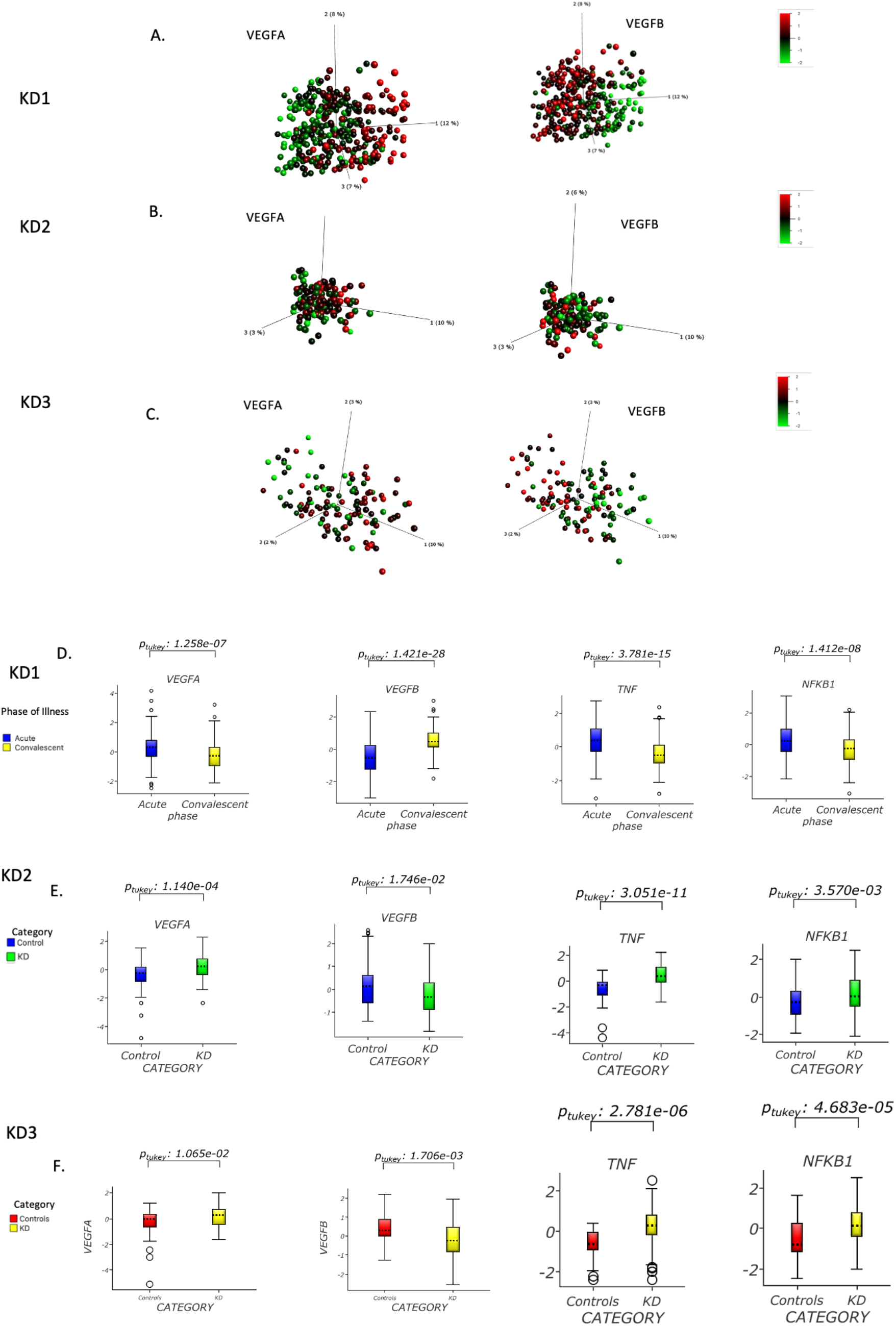
Angiogenesis related Gene-Expression in KD datasets. Kawasaki Dataset PCA heat map gene expression and box plots shown, KD1 (Figure **3A** &**B**), KD2 (Figure **3C** & **D**) and KD3 (Figure **3E** & **F**). For KD1 acute versus convalescent samples, KD2 and KD3 (KD versus control samples), there is a significant difference with downregulation of VEGFB, TNF and NFKB1 gene expression. For KD1 acute versus convalescent samples, KD2 and KD3 (KD versus control samples), there is significant upregulation in VEGFA gene expression.

### Comparing Response to IVIG Treatment (KD8 and KD9)

KD8 was analyzed to compare responders (R) versus non-responders (NR) to IVIG treatment (Figure 5D Supplement). For VEGF-A, VEGF-B, TNF, and NFKB1 gene expression there was no significant difference.

For KD9 R versus NR changes VEGF-A and VEGF-B gene expression are described before and after IVIG treatment (Figure 4). In KD9 there was no evidence of falls in NFKB1 and TNF gene expression. In KD9 both R and NR showed a significant fall in VEGF-A gene expression and a significant increase in VEGF-B gene expression. Also for KD9 prior to IVIG treatment, R had VEGF-B gene expression similar to controls, whereas for NR gene expression was significantly less compared to controls.

**Figure 4.**
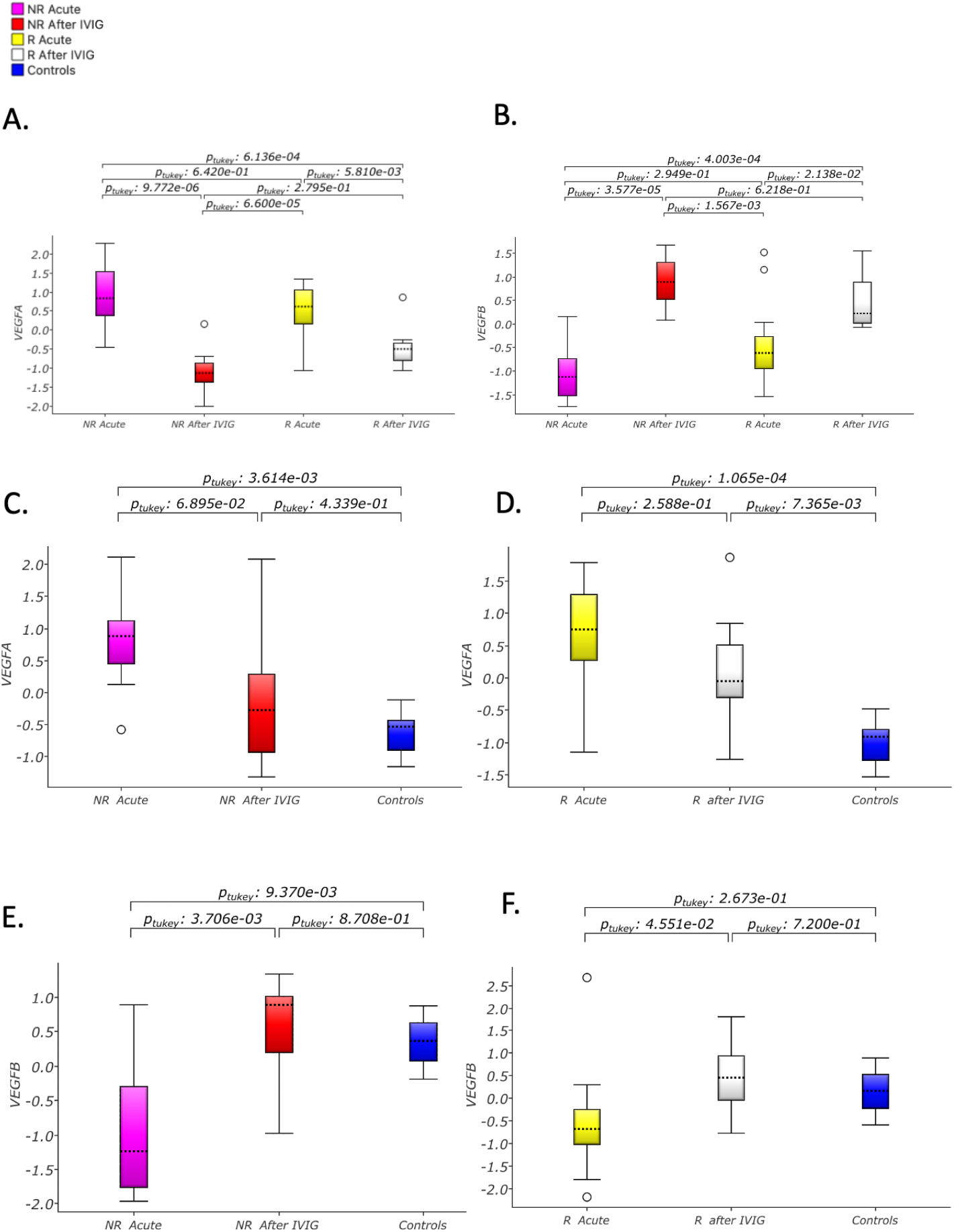
VEGF A and VEGFB Transcript patterns in KD patients with resistance versus responders to intravenous immunoglobulin treatment. KD9 Dataset (GSE18606) consisted of whole blood samples from 9 healthy age-appropriate controls, 8 Intravenous Immunoglobulin (IVIG) Non Responding (**NR**) and 12 IVIG Responding (**R**) patients. Non Responders before and after IVIG were compared to Responders before and after IVIG (**3A-B**). VEGFA and VEGFB gene expression compared to responders and non responders at similar respective stages of the analysis, pre and post IVIG, was no different. Non responders in the Acute phase are compared against Non responders after IVIG and controls (**3C-F**), similarly R is compared after IVIG and against controls. Responders to IVIG did not show a significant fall in VEGFA gene expresson before and after treatment. This is in contrast to Non responders who showed a significant fall in VEGFA gene expression. For both responders and non responders VEGFA gene expression is significantly different compared to controls before treatment, being similar to controls after IVIG treatment. VEGFB gene expression for both responders and non responders significantly increases after IVIG treatment. VEGFB gene expression is significantly down-regulated for non responders compared to controls, whereas for responders there is no difference before treatment. After treatment both responders and non responders show VEGFB gene expression that is similar to that of controls. When comparing the R and NR’s at their respective stages there was no significant difference in NFKB1 and TNF gene expression.

## DISCUSSION

This paper aimed to explore the role of VEGF in KD through VEGF subtypes A and B gene expression using Microarray and RNA-seq datasets. Firstly, gene enrichment of gene expression data from human coronary artery tissue of KD was analyzed using Rowley et al.’s original dataset {Rowley, 2015 #2394}. Here, VEGF-B gene expression was found to be down-regulated in coronary artery tissue of children with KD. However, TNF, NFKB1, and VEGF-A gene expression were not significantly different between KD cases and controls. One interpretation of this is that inflammation has subsided by the time of tissue harvest, potentially indicating that the Rowley et al. coronary tissue dataset depicts an end-stage process. These findings were consistent with Rowley et al.’s original study confirming that several cytokines and growth factors, such as VEGF-A, were not differentially expressed {Rowley, 2015 #2394}. However, gene expression suggests no change in inflammatory cytokines, enriched pathways related to innate and adaptive immune responses, cell signaling, and cytokine production. Further, gene enrichment analysis of the Rowley dataset elicited leukocyte-mediated immunity, migration dynamics, and adhesion to vascular endothelium. These are an essential part of coronary artery pathogenesis. This innate KD response is consistent with the work of Huang et al., who compared acute and convalescent cases of KD {Hoang, 2014 2320}. Thereby demonstrating the persistence of immunological pathway information from the post coronary artery harvest transcript can provide information about past immunopathogenesis.

The GSEA also confirmed gene pathways of significance on the Rowley Coronary Artery data. GO pathways were demonstrated in relation to the innate immune system and white cell dynamics (tethering or rolling). The latter, affecting white cells, was also shown by KEGG enrichment, evidencing leucocyte-endothelial interaction. Pathways related to NF-kB signaling and leukocyte extravasation signaling were also noted. The GO IL-6 and IL-8 pathways in KD4 are in keeping with the increase in levels of the same interleukins reported by Lin et al. in the first week of KD illness {Lin, 1992 #3222}. Also, in the first week of KD, Lin et al. found that IL-6 and IL-8 levels differentiated patients with and without coronary artery aneurysms. Further, in the analysis of KD4, no difference in VEGF-A and TNF, NFKB1 compared to controls was noted. However, this was in contrast to the datasets from the large multi-center studies (KD1, KD2, KD3), where TNF and NFKB1 were elevated in acute KD. However, in these studies, a significant decrease in TNF and NFKB1 gene expression was noted in the convalescent phase. Suggesting that Rowley’s data set is indeed representative of an end-stage process. The changing pattern of VEGF-A gene expression was also noted in our analysis of Fury’s original data {Fury, 2010 #3147}. Fury et al. mentioned the importance of NFKB signaling in KD. Also, showing that the CEACAM1 gene expression could differentiate between responders and non-responders to intravenous immunoglobulin therapy for KD. They suggested that CEACAM1 protein was necessary due to its relationship to the VEGF pathway. Therefore a pro-inflammatory phase evidenced by an elevated TNF and NFKB1 response is consistent with elevated VEGF-A, all according to gene expression patterns.

Three-dimensional PCA plots generated in two dimensions provided insights into understanding VEGF-A/B associations. This was applied to three landmark KD transcriptomic studies showing VEGF-A upregulation and VEGF-B downregulation. Also, one study suggested a temporal transition in acute versus convalescent in terms of VEGF-A/B gene expression. From VEGF-A upregulation and VEGF-B downregulation to VEGF-A downregulation and VEGF-B upregulation. Suggesting inversely related VEGF-A and VEGF-B gene expression changes. Hamamichi et al. showed VEGF levels to be significantly increased in acute KD and elevated maximally for two weeks after the onset of coronary artery lesions {Hamamichi, 2001 #2357}. Further, they illustrated a temporal difference in VEGF expression in neutrophils versus peripheral blood mononuclear cells (PBMCs). Thereby suggesting that neutrophil-derived VEGF could be part of an early response leading to coronary artery abnormalities. In our analysis, increased coronary artery VEGF-B gene expression was noted in the terminal phase of KD by Rowley et al.’s end-stage KD data with no difference in VEGF-A. However, a consistent pattern of VEGF-A/VEGF-B changes was noted when analyzing the large KD datasets. For example, our acute versus convalescent data analysis suggested an inverse temporal relationship between VEGF-A and VEGF-B. Jin et al. noted the counterregulatory role of VEGF-A and VEGF-B in adipose tissue differentiation {Jin, 2018 #2379}. VEGF-B is a homolog of VEGF-A {Arjunan, 2018 #3149} and this structural similarity may have temporal physiological implications. For example, we know that a differential affinity for the VEGF receptors, with VEGF-B having a stronger affinity for VEGR-2 compared to VEGF-A exists. Also, functional differences between VEGF-A and VEGF-B exist because VEGF-A has direct pro-angiogenic effects. In contrast, VEGF-B, though not directly angiogenic, could have a role in cell survival {Li, 2009 #2376}. The research could point to an intricate mechanism linking VEGF-A and VEGF-B in coronary artery disease associated with MISC and KD.

The role of VEGF-B in KD in cell survival and its relationship to VEGF-A has not been outlined in prior works. Against controls, VEGF-B was shown to increase gene expression, acting as a coronary growth factor in transgenic rats {Bry, 2010 #2381}. Moreover, Bry et al. also showed VEGF-B to have a counter role to VEGF-A, given that VEGF-B does not induce inflammation, cause a vascular leak, or angiogenesis. Additionally, VEGF-B and not VEGF-A antibodies inhibited coronary artery development {Tomanek, 2006 #2383}. Based on these facts, a vasculogenic role for VEGF-B is implicated {Lal, 2018 #2900}. Further, gene-targeted VEGF-B mice versus wild-type (WT) mice developed smaller hearts and dysfunctional coronary vasculature in the first postnatal month {Bellomo, 2000 #2382}. Also, VEGF-B gene therapy was shown to prevent doxorubicin cardiotoxicity through adenoviral delivery of a vector expressing VEGF-B{Rasanen, 2016 #2384}. They are evidencing the protective role of VEGF-B. Also, VEGF-B expression was associated with an improvement in endothelial function. VEGF-B may have a role as a safeguarding molecule for endothelial cells, smooth muscle, and pericytes{Li, 2009 #3152}{Zhang, 2009 #3157}. It is the most abundant in the heart and essential for cardiac blood vessel survival. Therefore a possible explanation for the inverse relationship between VEGFA and VEGFB is that VEGF-B is cardio-protective, countering the pro-inflammatory effect of VEGF-A in KD.

Based on our understanding of VEGF-A/B, with the possibility of a dual relationship between the two molecules, this could be exploited from a treatment perspective. Using knowledge of VEGF-related mechanisms through the study of tumors, various therapeutic options may be available {Korpanty, 2010 #2393}. A comparison of the effect of IVIG treatment on gene expression in KD was undertaken, showing no change in gene expression (study KD8). KD9 showed a fall in VEGFA expression for both R and NR, which was not elicited in dataset KD8. In Study KD9, VEGFB increased significantly for both NR and R. Therefore, the theme of a falling VEGFA and rising VEGFB was repeated in this one KD IVIG study, suggesting the relationship between VEGFA and VEGFB requires further exploration.

A limitation of the presented analysis concerns the fact that different studies cannot standardize for covariates such as age and sampling time-points, pre or post-treatment. Such variables are essential factors requiring consideration in gene expression studies. However, a cross-platform or meta-analysis was beyond the scope of this paper. Historically, KD studies have been primarily cross-sectional in design; thus, taking temporal relationships into account is challenging. A longitudinal approach could be more beneficial for future research wherein including additional clinical parameters would be desirable. Also, in KD research, another limitation is the inability to follow clinical changes in the coronary arteries closely. However, clinical in-situ gene expression data studies from coronary arteries are not technically possible during KD. Engaging with the Rowley et al. coronary artery dataset was an attempt to understand end-stage disease{Rowley, 2015 #2394}.

## Conclusion

A VEGF-A/VEGF-B mRNA gene expression was explored in KD datasets chosen from a systematic search. VEGF-A is a known angiogenic molecule with proinflammatory effects whereas the role of VEGF-B has been less defined in the clinical literature. Subsequent transcriptomic analysis across several studies provided a novel cross validation of temporal patterns in VEGF-A and VEGF-B gene expression. This suggests that VEGF-A and VEGF-B have possibly an intricate inverse relationship. On this basis a dual therapeutic strategy, enhancing VEGF-B whilst minimising VEGF-A effects could be a possible advancement over current KD clinical strategies. VEGF-A/B dual relationship may have implications beyond the field of sepsis, given the extensive study of VEGF-A in tumours. This study we believe surpasses the impasse in understanding the role of VEGF-B in disease pathogenesis. Investigating this dual relationship may be more productive in the future with implications for KD associated coronary artery disease prevention. Therefore, further research, exploring the pathophysiological relationship between VEGF-A and VEGF-B in KD is suggested.

## Data Availability

Publicly available on the GEO NCBI Webserver at https://www.ncbi.nlm.nih.gov/geo/ with Accession Numbers GSE63881, GSE73461, GSE68004, GSE64486, GSE16797, GSE15297, GSE18606, GSE48498, GSE109351

## Acknowledgments

We are extremely grateful to the Sheffield Institute of Translational Neuroscience for technical advice (SITRaN), in particular Dr. Paul Health. To Ege Ulgen for assistance with the data.

## Author Contributions

Conceived and designed the experiments: AR

Performed the experiments: AR

Analyzed the data: AR, MT

Contributed analysis, methods, and tools: AR, MT, BSB

Wrote the first draft of the paper: AR

Revised critically for importance and intellectual content: AR, MT, PK, ZAM, JS, RK, BSB, AS, DC, RM, NQ, GB, SAZ, PO, WZ, MAZ, HS, AAK, MRN, GS, SH, MT, AA, AH

## Research and Ethics Committee

The paper was waived from review by the hospital Central Scientific Committee and the Research and Ethics Committee given the secondary analysis of data from primary studies.

