## Supplement Table 3 for "VEGF subtype A and B Gene Expression, Clues to a Temporal Signature in Kawasaki Disease, Implications for Coronary Pathogenesis through a Secondary analysis of Clinical Datasets"

**Table 3 (supplement).** Gene Expression analysis of KD associated coronary arteritis.

A.

| Pathway | nGenes | Pathway Genes | Fold Enrichment | Enrichment FDR |
|---|---|---|---|---|
| Regulation of antigen receptor-mediated signaling pathway | 12 | 69 | 11.55837242 | 3.79E-08 |
| B cell receptor signaling pathway | 11 | 67 | 10.91144859 | 3.56E-07 |
| Positive regulation of interferon-gamma production | 12 | 90 | 8.861418853 | 7.88E-07 |
| Regulation of interferon-gamma production | 16 | 136 | 7.818898988 | 2.18E-08 |
| Antigen receptor-mediated signaling pathway | 33 | 326 | 6.727610939 | 2.22E-15 |
| Positive regulation of T cell activation | 27 | 280 | 6.408704706 | 2.95E-12 |
| T cell differentiation | 27 | 281 | 6.385897928 | 3.11E-12 |
| Positive regulation of leukocyte cell-cell adhesion | 29 | 307 | 6.278041044 | 6.42E-13 |
| T cell receptor signaling pathway | 26 | 276 | 6.260785059 | 1.34E-11 |
| T cell activation | 51 | 574 | 5.905039567 | 1.94E-21 |
| Leukocyte cell-cell adhesion | 39 | 445 | 5.824640482 | 3.24E-16 |
| Regulation of T cell activation | 36 | 412 | 5.807240511 | 5.70E-15 |
| Lymphocyte differentiation | 35 | 406 | 5.729365638 | 2.10E-14 |
| Regulation of leukocyte cell-cell adhesion | 34 | 408 | 5.538386783 | 1.32E-13 |
| Positive regulation of cell activation | 34 | 427 | 5.291948027 | 4.65E-13 |
| Lymphocyte activation | 63 | 825 | 5.075176252 | 2.91E-23 |
| Lymphocyte proliferation | 26 | 357 | 4.840270802 | 3.78E-09 |
| Adaptive immune response | 47 | 646 | 4.835371743 | 1.71E-16 |
| Regulation of leukocyte activation | 44 | 629 | 4.649075074 | 5.70E-15 |
| Positive regulation of immune response | 53 | 830 | 4.243872282 | 2.08E-16 |
| Positive regulation of immune system process | 71 | 1190 | 3.965298773 | 9.67E-21 |
| Regulation of cell adhesion | 49 | 837 | 3.890766342 | 7.56E-14 |
| Leukocyte migration | 27 | 502 | 3.57457633 | 9.59E-07 |
| Regulation of immune response | 79 | 1488 | 3.528488354 | 2.13E-20 |
| Positive regulation of cytokine production | 33 | 628 | 3.492358545 | 4.37E-08 |
| Cytokine production | 45 | 937 | 3.191813087 | 6.14E-10 |
| Innate immune response | 49 | 1258 | 2.58868953 | 7.76E-08 |
| Immune effector process | 47 | 1245 | 2.50895594 | 4.60E-07 |
| Response to biotic stimulus | 70 | 1905 | 2.442123306 | 1.96E-10 |
| Defense response to other organism | 55 | 1499 | 2.438515862 | 5.74E-08 |

B.

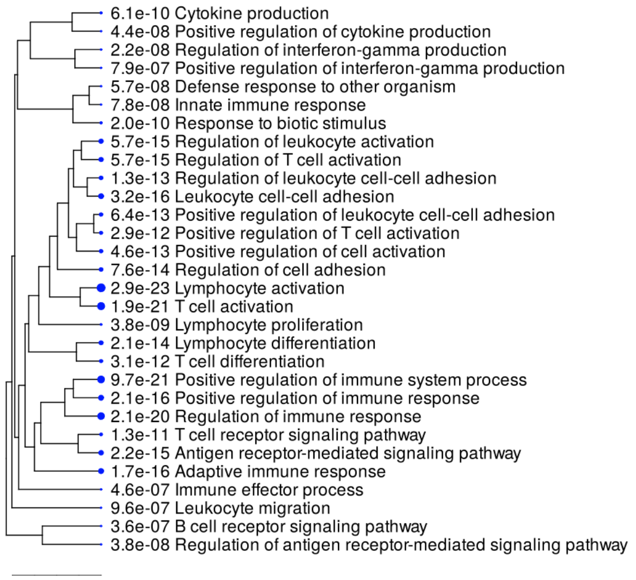


C.

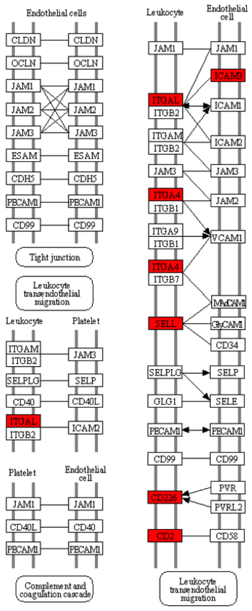


**Table 3 (supplement).** From KD4, t-Test children with Kawasaki Disease versus controls from coronary artery tissue (p=0.001, q=0.12) elicited 527 genes which then underwent enrichment and the GO network generated on the ShinyGo platform. The enriched GO terms are shown **(A).** The GO terms underwent hierarchical tree clustering showing the correlation amongst the significantly enriched pathways **(B).** Enriched genes were also mapped onto the KEGG database, here depicted as a KEGG graph using Pathview **(C)** showing genes enriched in red on the leucocyte side and endothelium.
