## Supplementary material for "VEGF subtype A and B Gene Expression, Clues to a Temporal Signature in Kawasaki Disease, Implications for Coronary Pathogenesis through a Secondary analysis of Clinical Datasets": Table 2

**Table 2.** Coronary Artery Gene Set Expression analysis (GSEA)

| Pathway | NES | p | q |
|---|---|---|---|
| Biocarta Cytokine Production | 1.71 | 0.006 | 0.11 |
| GO B Cell Mediated Immunity | 2.2 | 0 | 0.001 |
| GO Cellular Extravasation | 1.67 | 0.03 | 0.12 |
| GO Cytokine Metabolic Process | 1.75 | 0.04 | 0.09 |
| GO Defence Response to Bacterium | 2.06 | 0 | 0.003 |
| GO Huoral Immune Response | 2.08 | 0 | 0.002 |
| GO Immunoglobulin Production | 2.07 | 0 | 0.002 |
| GO Leucocyte Adhesion to Vascular Endothelial Cell | 1.72 | 0.04 | 0.1 |
| GO Leucocyte Tethering or Rolling | 1.76 | 0.006 | 0.1 |
| GO Positive Regulation of Cell Cell Adhesion | 1.7 | 0.03 | 0.11 |
| GO Positive Regulation of Cytokine Production | 1.59 | 0.049 | 0.15 |
| GO Positive Regulation of Interferon Gamma Production | 1.84 | 0.008 | 0.05 |
| GO Positive Regulation Interleukin 1 Production | 1.71 | 0.04 | 0.1 |
| GO Positive Regulation Interleukin 6 Production | 1.74 | 0.04 | 0.1 |
| GO Positive Regulation Interleukin 8 Production | 1.62 | 0.03 | 0.14 |
| GO Tumor Necrosis Factor Receptor Binding | 1.62 | 0.03 | 0.15 |

**Table 2.** Coronary artery dataset KD4 (GSE 64486) underwent GSEA of Cases versus controls. Variables were identified according to the averaging of their Genes Symbols. A two group comparison was undertaken $p<0.05$ and $q <0.25$. Elicited GO pathways are shown. Gene set used was TNF.gmt for GSEA.
