## Supplementary material for "VEGF subtype A and B Gene Expression, Clues to a Temporal Signature in Kawasaki Disease, Implications for Coronary Pathogenesis through a Secondary analysis of Clinical Datasets": Table 1

### TABLE 1: Summary of KD Clinical Studies Analysed

| Accession number* | Datatype | Platform & Datatype | Institution | Research Facility | Dataset Name | STUDY DESIGN | n | Paper Reference |
|---|---|---|---|---|---|---|---|---|
| GSE63881 | Microarray | Illumina Human HT-12 V4.0 beadchip | Rady Childrens Hospital San Diego | Genome Institute of Singapore | KD1 | Case-controlled study. KD Acute versus convalescent samples. | 146 | {Hoang, 2014 2320} |
| GSE73461 | Microarray | Illumina Human HT-12 V3.0 beadchip | Pediatric centers in the UK, Spain, Netherlands and the USA | Imperial College UK | KD2 | Case-controlled study | 616 | {Wright, 2018 #2323} |
| GSE68004 | Microarray | Illumina Human HT-12 V4.0 beadchip | Nationwide Children's Hospital in Columbus<br>Children's Medical Center Dallas<br>Rady Children's Hospital in San Diego | BIIR Lab Dallas USA | KD3** | KD children were enrolled and sampling prior to IVIG and then testing repeated in the convalescent phase of the illness. | 162 | {Jaggi, 2018 #2321} |
| GSE64486*** | RNA-Seq | Illumina HiSeq 2000 | Children's Hospital Chicago<br>USA-wide cases | Barts & The London School of Medicine UK | KD4 | Case-controlled study. Coronary artery tissue from KD (n=8) and childhood controls (n=7) | 15 | {Rowley, 2015 #2394} |
| GSE16797 | Microarray | Affymetrix Human Genome U133 Plus 2.0 Array | Kitasato University Hospital<br>Ebina General Hospital | Kitasato University Hospital or Ebina General Hospital | KD5 | The Egami scoring system was used to predict the IVIG-responsive (Group A; n = 6) and IVIG-resistant groups (Group B) before starting the commencement of treatment using. Also random allocation of the following groups was undertaken, single-IVIG treatment (Group B1; n = 6), combined therapy group (Group B2; n = 5) and IVIG-plus-methylprednisolone (IVMP). | 17 | {Ogata, 2009 #3140} |
| GSE15297 | SMD Print_853 | Stanford methodology using Ambion amplification kit followed by fluorescent labelling | Rady Children's Hospital San Diego and<br>Children's Hospital Boston | University of California, San Diego Children's Hospital Boston and Stanford University | KD6 | Case-controlled study.<br>23 Patients with KD, 18 febrile controls and 10 non febrile controls | 41 | {Popper, 2009 #3141} |
| GSE109351 | Microarray | Affymetrix Human Transcriptome Array 2.0 | Kaohsiung Chang Gung Memorial Hospital<br>Children's Hospital<br>Kaohsiung | Department of Medical Research<br>Genomics and Proteomics Core Lab<br>Kaohsiung | KD7 | Case-controlled study. Comparison of heathy controls versus acute and convalescent phase KD. There groups of RNA samples were analysed. 3 pooled acute-phase KD RNA samples and 3 convalescent-phase KD RNA samples and 3 pooled healthy control RNA samples. | 9 | {Guo, 2022 #3145} |
| GSE48498 | Microarray | Affymetrix Human Genome U133 Plus 2.0 Array | Kitasato University Hospital | Agilent TechnologiesSanta Clara, California, USA | KD8 | Pre- and post-treatment effects were analysed in IFX-responsive patients (n=8) and IVIG-responsive patients (n=6), resulting in a sample number of 28. | 14 | {Ogihara, 2014 #3148} |
| GSE18606 | Microarray | Agilent G4112F Human Whole Oligo Array | Rady Children's Hospital San Diego | Baylor Institute for Immunology Research NIAID Cooperative Centers Luminex Core Facility | KD9 | Comparing responders to IVIG versus non responders. The following groups were analysed. 9 healthy age-appropriate controls, 12 IVIG-responders and 8 IVIG non-responders at acute and convalescent stages, resulting in 48 whole blood samples. | 29 | {Fury, 2010 #3147} |

* NCBI GEO Accession Numer or NCBI Bioproject ID
** On Trimmed Mean of the M-Values (TMM) normalisation there was filtering out of certain gene probes
HB = Whole Blood
CA = Coronary Artery
IVIG = Intravenous Immunoglobulin
IFIX = Infliximab
