## Supplement Figure 5 for "VEGF subtype A and B Gene Expression, Clues to a Temporal Signature in Kawasaki Disease, Implications for Coronary Pathogenesis through a Secondary analysis of Clinical Datasets"

**Supplementary Figure 5.** VEGF A and VEGFB Transcript patterns in KD patients with resistance versus
responders to intravenous immunoglobulin treatment.

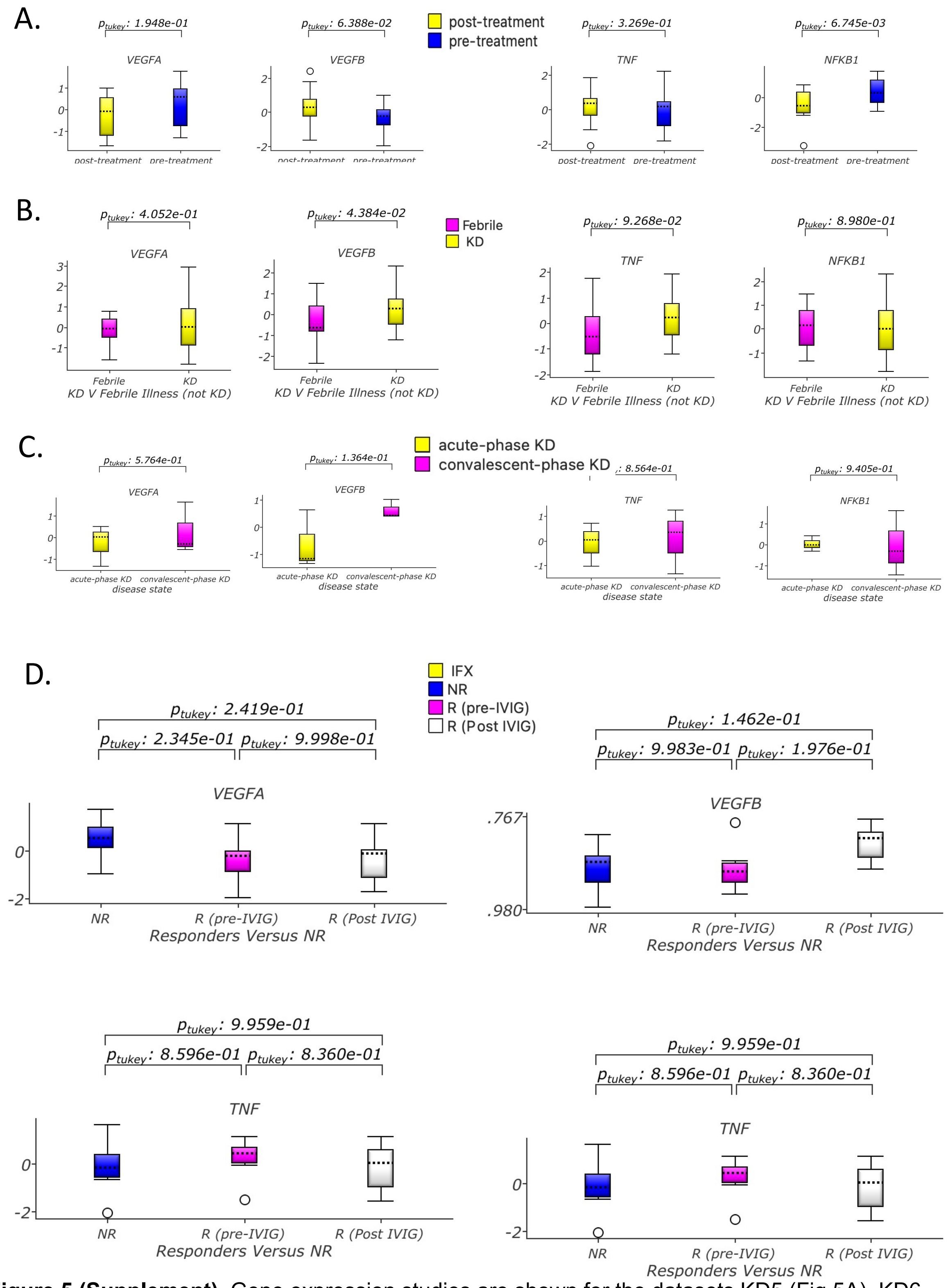


**Figure 5 (Supplement).** Gene expression studies are shown for the datasets KD5 (Fig.5A), KD6 (Fig.5B), KD7 (Fig5C) and KD8
(Fig.5D). KD8 R pre and post IVIG administration are shown agains NR.
